# A Mixed Methods Study Evaluating Acceptability of a Daily COVID-19 Testing Regimen with a Mobile-App Connected, At-Home, Rapid Antigen Test: Implications for Current and Future Pandemics

**DOI:** 10.1101/2022.04.18.22273982

**Authors:** Nadia Nguyen, Benjamin Lane, Sangwon Lee, Sharon Lipsky Gorman, Yumeng Wu, Alicia Li, Helen Lu, Noemie Elhadad, Michael Yin, Kathrine Meyers

## Abstract

**Background:** Widespread use of at-home rapid COVID-19 antigen tests has been proposed as an important public health intervention to interrupt chains of transmission. Antigen tests may be preferred over PCR because they provide on-demand results for relatively low cost and can identify people when they are most likely to be infectious, particularly when used daily. Yet the extent to which a frequent antigen testing intervention will result in a positive public health impact for COVID-19 will depend on high acceptability and high adherence to such regimens.

**Methods:** We conducted a mixed-methods study assessing acceptability of and adherence to a daily at-home mobile-app connected rapid antigen testing regimen among employees of a US-based media company. Acceptability was assessed across seven domains of the Theoretical Framework of Acceptability.

**Results:** Among 31 study participants, acceptability of the daily testing intervention was generally high, with participants reporting high perceived effectiveness, intervention coherence, and self-efficacy; positive affective attitude; acceptable degree of burden and opportunity cost; and assessing the intervention as ethical. 71% reported a preference to test daily using an at-home antigen test than weekly employment-based PCR. Mean adherence to the 21-day testing regimen was 88% with 43% of participants achieving 100% adherence, 48% testing at least every other day, and 10% testing less than every other day.

**Conclusions:** Despite overall high acceptability and adherence, we identified three implementation challenges that must be addressed for frequent serial testing for COVID-19 to be implemented at scale and have a positive public health impact. First, users need guidance on how and when to adapt testing frequencies to different epidemiological conditions. Second, users and institutions need guidelines for how to safely store and share test results. Third, implementation of serial testing strategies must prioritize health equity and protect those most vulnerable to COVID-19.

## 1.0 Introduction

The use of at-home rapid COVID-19 antigen tests has been identified as an important public health intervention because such tests provide on-demand results for relatively low cost and can identify people when they are most likely to be infectious. Although antigen tests are not as sensitive as molecular tests like real-time PCR for detecting COVID-19, this limitation can be overcome by testing more frequently, including as often as daily. There is growing evidence from both modeling (1,2) and real-world studies (3–6) that antigen tests used frequently perform as well, and in some cases better at controlling the spread of COVID-19, than PCR administered less frequently.

Although antigen tests were developed relatively early in the pandemic, with the first at-home antigen test gaining Food and Drug Administration (FDA) approval for use in the United States (US) in December 2020 (Figure 1), they have been unavailable and underutilized for much of the pandemic in favor of more sensitive but also more expensive and time-consuming lab-based PCR tests (7). In contrast, at-home antigen testing is a cornerstone in COVID-19 control measures in many European countries, including the UK and Germany, where such tests have been provided for free or at very low cost and have been widely used (7–9). Recognizing the important role of at-home antigen tests, the Biden administration has taken a number of steps to significantly increase the availability and use of at-home antigen tests, including scaling up production of such tests; investing $2 billion to distribute free tests throughout the community; selling tests at cost; mailing free tests to households; and mandating tests be reimbursed through insurers (10–12). These steps have the potential to dramatically increase the availability of and access to at-home antigen in the US.

**Figure.**
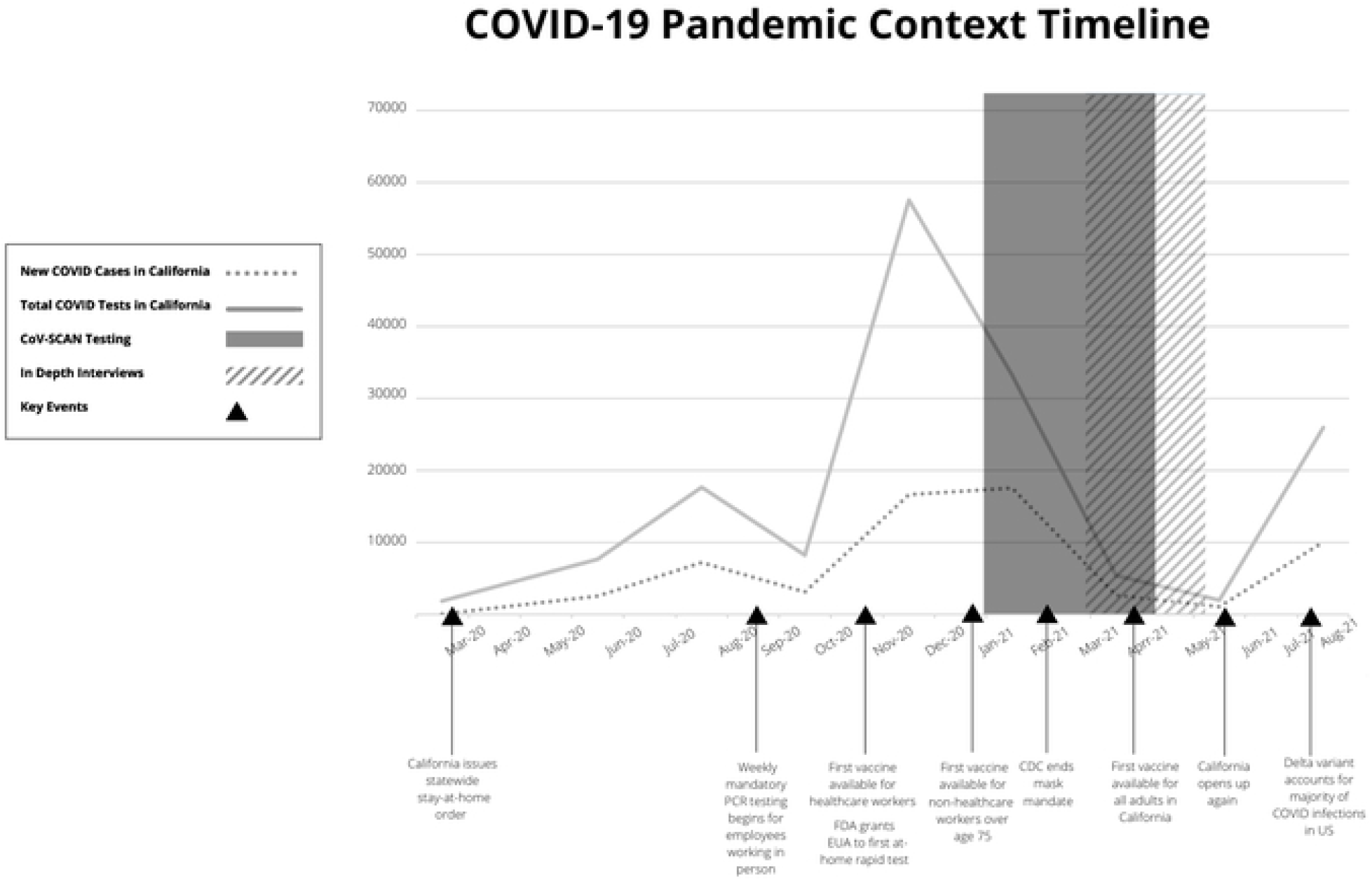

Yet the extent to which a massive influx of at-home antigen tests to the American public will result in a positive public health impact will depend on whether and how the tests are used. To date, there is limited real-world data on the acceptability of at-home antigen testing, particularly when used serially, where they have been shown to be most effective. Previous implementation studies of numerous evidence-based interventions, particularly those requiring sustained participation and adherence, indicate that acceptability is critical to intervention uptake and success (13–15). Even if at-home antigen tests become widely available, their public health impact will be limited if frequent testing is not acceptable and adopted at scale. To address this knowledge gap, we conducted a mixed-methods study informed by Sekhon’s Theoretical Framework of Acceptability (TFA) (16) to evaluate the acceptability of and adherence to a daily testing regimen among employees of a large media company in the US.

## 2.0 Methods

### 2.1 Research Design

We conducted a mixed-methods observational study from February-June 2021 to examine factors that influenced the acceptability of and adherence to a serial testing regimen using a self-administered at-home rapid antigen test (“at-home antigen test”) in a workplace setting among employees at two divisions of The Walt Disney Company (“Disney”), a media company based in Southern California in the US. This mixed-methods acceptability study was nested within a larger parent study (N=93) testing the effectiveness of a daily testing regimen using an at-home antigen-test against weekly testing with PCR at Disney. Participants in the parent study were mailed at-home COVID-19 antigen test kits to their home address and asked to self-test daily for 21 days and, as a condition of their employment, were also concurrently testing at least weekly for COVID-19 using PCR. In our nested acceptability study, after providing written informed consent, participants were asked to complete a one-time quantitative survey (N=31), and a subset of participants contributed a one-time qualitative in-depth interview (N=15) at the conclusion of 21 days of daily testing. Participants had a choice to complete both the quantitative survey and qualitative interview or just the survey. Participants who completed the survey were entered into a drawing for a $100 gift card, and participants who completed the interview received a $25 gift card. All study procedures were reviewed and approved by the Columbia University Irving Medical Center’s Institutional Review Board.

### 2.2 Recruitment and Study Population

Participants in the acceptability study were informed of the goals of the study and recruited from the parent study via email. To be eligible for the parent study, participants were 18 years of age or older; owned a smartphone; understood and read English; had not been diagnosed with COVID-19 in the past 90 days; had not received any COVID-19 vaccine doses at enrollment; and were willing to share their weekly PCR results. There were no additional inclusion criteria for the acceptability study aside from willingness to complete the quantitative survey and being at least 14 days into the parent. COVID-19 vaccinated employees were excluded because at the time of recruitment, vaccination was believed to provide strong protection against breakthrough infections.

### 2.3 At-Home Rapid Antigen Test and Mobile App

Participants used an investigational at-home antigen test that is currently under review with the FDA for Emergency Use Authorization. Test results were available to participants after 15 minutes (for test details, see Appendix A) The test also included a paired smart phone mobile application (mobile app) that had the following functions: provided step-by-step testing instructions; captured image of test result and automatically interpret result (positive, negative, invalid; securely store test result history; and transmit daily test result to the study team.

### 2.4 Data Collection

#### 2.4.1 Theoretical Framework

The quantitative survey and in-depth interview questions were informed by Sehkon’s Theoretical Framework of Acceptability (TFA) (16). The TFA defines intervention acceptability as a multi-faceted construct that reflects the extent to which people consider the health care intervention to be appropriate, based on anticipated or experienced cognitive and emotional responses to the intervention. The TFA consists of seven constructs which we adapted to measure the acceptability of the intervention: affective attitude, burden, perceived effectiveness, ethicality, intervention coherence, opportunity costs, and self-efficacy (Table 3).

#### 2.4.2 Adherence to Daily Testing Regimen

Participants submitted daily test results to the study team by scanning a picture of the test cassette using the mobile app.

#### 2.4.3 Quantitative Survey

Each of the TFA’s seven constructs of acceptability was evaluated by one to three statements which participants rated on a five-point Likert scale (strongly disagree=1 to strongly agree=5). As a global measure of acceptability, participants were also asked whether, if given a choice, they would prefer to test for COVID-19 using the current PCR regimen or test daily with an at-home antigen test. We measured sociodemographic factors, including age, race/ethnicity, sex, education, living environment, and financial stability. Participants self-administered the 20-minute survey using REDCap, a secure web platform for administering online surveys.

#### 2.4.4 In-Depth Interview

We conducted in-depth interviews with participants to more deeply understand their current and previous COVID-19 testing experiences and the acceptability of the daily COVID-19 testing regimen. The interview guide was designed to assess the seven constructs in the TFA. Interviews were approximately 60 minutes long and conducted over Zoom by a team of four researchers experienced in qualitative interviewing (NN, BL, KM, and YW). Of the four researchers: two identified as Asian cis-gendered females, 1 White cis-gendered female, and 1 White cis-gendered male; all four lived in New York City. All four were employed by Columbia University, which made rapid turnaround PCR testing available as needed in addition to random surveillance testing, and all were also using at-home antigen tests in their personal lives.

Interviews were summarized in debrief forms immediately after completing the interviews and were audiorecorded and transcribed by a paid transcription service.

### 2.5 Data Analysis

#### 2.5.1 Quantitative analysis

We summarized participant characteristics by estimating mean, standard deviation, frequency and proportion for all categorical and continuous sociodemographic and health factor variables, respectively. Additionally, we estimated the mean, standard deviation, median, and interquartile range (IQR) for each TFA construct measure and also estimated the frequency and proportion of participants who stated that they strongly agreed or agreed with each statement. We calculated overall adherence by taking the number of submitted test results and dividing it by the 21 days of the study. We categorized adherence into three groups and estimated the number and proportion of participants per group: 100% (tested daily), 50%-99% (tested at least every other day), and <50% (tested less than every other day). Finally, we calculated the frequency and proportion of participants who stated they would prefer testing daily with a rapid antigen test versus their current PCR regimen if given the choice.

#### 2.5.2 Qualitative analysis

Interview transcripts were analyzed by a team of three researchers in Dedoose using Template Analysis, which is appropriate for studies using a priori themes and exploring perspectives of different groups in an organizational context (17). We initiated data analysis with a top-down approach, using the seven constructs in the TFA to create an organizational template. One member of the research team created a preliminary codebook by reading through interview debrief forms and identifying subconstructs within each of the TFA constructs. With the seven TFA constructs listed as parent codes and the subconstructs as child codes, the initial codebook was applied to two interview transcripts in Dedoose. A second researcher then applied the preliminary codebook to two more interviews and the codebook was further discussed and refined. All three coders then re-coded the initial four interviews using the finalized codebook to maximize reliability across coders. The three coders then individually coded the remaining 11 interviews, meeting on an ad hoc basis to resolve coding questions, and using memos to note emerging themes. The coders generated code reports for each TFA construct, identified key quotes for each construct and subconstruct, and held team meetings to discuss emergent themes that conceptualized how each construct either directly or indirectly influenced acceptability and adherence. Lastly, we created quote matrices for each construct and subconstruct using the identified key quotes organized by emergent themes.

## 3.0 Results

### 3.1 Participant characteristics

Of the 63 eligible Disney employees, we enrolled 31 participants who completed the online survey, of whom 15 also agreed to an in-depth interview between February-June 2021 (Table 1, Figure 1). The 15 interview participants were similar to the overall sample of the survey, with regard to sociodemographic and health factors and adherence.

### 3.2 Adherence to daily testing regimen

Mean adherence to the 21-day testing regimen was 88% with 43% of participants achieving 100% adherence, 48% achieving 50-99% adherence, and 10% achieving less than 50% adherence (Table 2).

### 3.3 Testing preference

Most participants (71%) reported that if given the choice, they would prefer to test for COVID-19 daily using an at-home antigen test than weekly using PCR, while 19% preferred PCR, and 10% preferred only testing when necessary (Table 2).

### 3.4 Acceptability of the daily testing regimen

#### 3.4.1 Affective Attitude

Participants reported overall positive feelings about the intervention though only 58% agreed with the statement “I like using the test every day” (see Table 3 for mean and median Likert scale scores). The overwhelming feeling reported by participants was comfort and peace of mind (Table 4, Affective attitude 1). Testing negative gave them needed reassurance to go about their daily lives and participants reported feeling “liberated” to engage in everyday activities without fear of infecting others (Affective attitude 2, 3). Participants also described using the tests after engaging in a “high risk” activity or potentially being exposed to COVID-19 (Affective attitude 3). Notably, a small number of participants described significant anxiety due to living with someone who was immunocompromised; for these participants, the ability to test daily or on demand provided significant mental health benefits (Affective attitude 4).

For some participants who struggled to incorporate testing into their daily routine or who had difficulty using the mobile app to capture and interpret their test result, daily testing was more burdensome. In these cases, participants reported feeling dread when they remembered they still had to test or frustration at being repeatedly unable to scan in a test result through the mobile app (Burden 6, Opportunity cost 5).

#### 3.4.2 Burden

More than half of participants disagreed with the statements “it is inconvenient to use the self-test every day” (61%) (Table 3). Most participants describing daily self-testing as very convenient compared to other testing options they experienced (e.g., large drive-through testing events early in March/April 2020 or mandatory testing offered by their employer). Even among participants who had access to weekly on-site testing at their workplace, most preferred the daily antigen test. These participants overwhelmingly described testing daily as fast and easy often noting that daily self-testing took less time overall than weekly PCR testing (Table 4, Burden 1); that they didn’t mind daily testing because they could test on their own schedule and get the results quickly (Burden 2) or incorporate self-testing into other established routines (e.g., timing the 15-minute waiting period with a shower) (Burden 3); and that they did not find the daily swabs to be invasive (Burden 4). While some participants described mild discomfort from the daily swabbing, this was not a major burden.

In the minority were participants who acknowledged that while daily at-home antigen testing was more convenient than weekly PCR testing, they still preferred testing using PCR because of the mental energy required to self-administer the test (Burden 5). Other participants described difficulty incorporating testing into their daily routine and reported testing fatigue (Burden 6).

#### 3.4.3 Self-efficacy

Participants reported that the self-test was easy to learn to use (90%) and results easy to understand (100%) (Table 3). However, only half of participants (55%) stated that they developed a habit of testing for COVID-19 in their daily routine, with experiences adhering to daily testing varying widely across participants. While some participants reported feeling initially overwhelmed by the testing steps, nearly all reported mastering the steps after a few days of testing (Table 4, Self-efficacy 1, 2). Factors that supported daily adherence included developing strategies to streamline the testing process (Self-efficacy 2); incorporating the test into their daily routine (Self-efficacy 3, 4); and placing the test kit in a prominent location (Self-efficacy 3). In contrast, participants reported that changes to their daily routine and weekends were the greatest barrier to daily adherence (Self-efficacy 5, 6).

#### 3.4.4 Opportunity Cost

Most participants strongly agreed with the statement: “It is easy for me to find a time to use the test every day” (68%) (Table 3). Nonetheless, time was often described as the biggest opportunity cost to testing daily (Table 4, Opportunity cost 1, 2). Some participants attempted to minimize the time cost by multi-tasking (Burden 3) or spreading out the time commitment by setting up their test kit the night before (Self-efficacy 3). However, others reported that efforts to multi-task resulted in more time spent testing because they would lead to mistakes like missing the results window during and having to repeat the testing process again (Opportunity cost 1, 2, 3). For other participants, access to strong internet and/or a good light source were additional opportunity costs to daily testing as the mobile app required a specific level of light to accurately interpret the test result (Opportunity cost 4, 5). Participants who struggled to upload a picture of their test result consequently described testing as a “daily hassle” and “frustrating”, negatively impacting their affective attitude about the testing regimen.

#### 3.4.5 Ethicality

Most participants agreed that it is ethical for an employer to require its employees to test regularly for COVID-19 (87%) (Table 3). However, a small minority believed that regular testing should be a personal choice (10%) and a significant portion of participants believe that sharing test results should be a personal choice (42%). Many participants articulated how working long hours near other coworkers necessitated the need for routine testing and saw mandatory testing as a practical approach to keeping the workplace safe (Table 4, Ethicality 1, 2). Specifically, many used Utilitarian arguments to justify mandatory testing, saying that the small loss of privacy and autonomy was justified for “the greater good” of workplace safety (Ethicality 3, 4, 5). Others went further, stating that routine testing helped them fulfil their personal responsibility to keep their workplace safe, noting that they “didn’t want to be transmitters” and that giving someone else COVID would be “an unacceptable thing” (Ethicality 6). In contrast, a small number of participants believed mandatory workplace testing was not ethical as it infringed upon individual liberties. While these participants were personally willing to test regularly through their employer, they believed that coercive tactics should not be used to keep the workplace safe (Ethicality 7).

While participants tended to feel strongly that requiring regular COVID-19 tests was either ethical or unethical, participants’ feelings about who should have access to test results and what this health information should be used for was more nuanced. Many agreed that test results should be reported to their employer to prevent the spread of infection should an employee test positive (Ethicality 8); however, some had concerns about who within their company should have access to results (Ethicality 9). Participants also had mixed feelings regarding whether COVID-19 test results should be shared with larger health organizations, such as state health departments or the Centers for Disease Control and Prevention (CDC). Most participants believed these were trusted entities and saw the benefit of sharing COVID-19 results for contract tracing, surveillance, and allocation of health services (Ethicality 10); however, this feeling was not universal, and some reported not trusting the CDC or worrying that the health department would limit their freedom if they tested positive (Ethicality 11, 12). Additionally, a small number of participants believed test results should stay within their employer due to concerns about health information privacy, including fears that their health information could be sold or compromised (Ethicality 13, 14). At the same time, many participants liked the idea of their test results being automatically reported to their employer or doctor through a mobile app, as this would eliminate the need for a third party to handle test results (Ethicality 15).

#### 3.4.6 Coherence

The majority of participants believed that testing for COVID-19 daily made sense to prevent the spread of COVID-19 at and beyond the workplace (77%) (Table 3). Support for daily testing was particularly strong for times when COVID-19 rates were increasing or high; in this context, regular testing was considered an effective and necessary strategy for monitoring infection rates and preventing COVID-19 transmission (Table 4, Coherence 1). Specifically, many participants believed the unique needs of the entertainment industry, which relies on non-replaceable people to fill specific roles, further necessitated a stricter approach to COVID-19 control to ensure that work would not be disrupted (Coherence 2). Many participants said they wish they had access to daily testing during the original peak of the pandemic in March and April of 2020 (Coherence 3).

While support for regular testing was near universal during times when risk of COVID-19 infection was perceived to be high (i.e., before vaccines were available, when rates of infection were still increasing), there was less agreement about when routine testing was no longer needed. Many participants thought it made sense to continue regular COVID-19 testing until there was more research available on the transmissibility of emerging COVID-19 variants and effectiveness of the COVID-19 vaccines (Coherence 5). Others preferred to listen to scientific recommendations from public health agencies to decide whether frequent testing made sense (Coherence 6). Some participants expressed increasing doubt that regular COVID-19 testing was still necessary given that vaccines were widely available to all adults and rates of COVID-19 in the workplace and surrounding community were very low (Figure 1). One participant in a managerial position used a cost benefit analysis to argue that frequent testing did not make sense from a financial perspective given the low positivity rates (Coherence 7). Many believed testing did not make sense for vaccinated populations, but that it may be an alternative for those who did not wish to become vaccinated (Coherence 8).

#### 3.4.7 Perceived effectiveness

Participants had high confidence in the effectiveness of a daily testing intervention with the majority reporting that they trusted the results of the at-home antigen test (81%) and nearly all expressing confidence that testing daily will keep the people they work with safe from COVID-19 (90%) (Table 3). Whether participants perceived the daily testing intervention would keep the workplace safe depended on three factors: 1) whether they trusted the test results, 2) whether they believed their co-workers would be willing and able to test daily, and 3) whether they believed frequent testing would introduce any unintended consequences. Although the antigen test used in the study had not yet received FDA approval and was only for use in research settings, nearly all participants reported trusting the results, citing trust in Columbia University (which helped develop the test in collaboration with a biotechnology company), trust in Disney (their employer) to provide a safe and effective test, and trust in experts/scientists (Table 4, Perceived effectiveness 1, 2). Other participants drew on previous experience using similar at-home tests (e.g., home pregnancy tests, glucose monitoring, maintaining swimming pool pH) as reasons for trusting the results (Perceived effectiveness 3). One participant noted that they trusted the daily antigen test result more than weekly PCR because the antigen test result reflected current COVID-19 status while the PCR result reflected a status from 24-48 hours ago and could give a false sense of security (Perceived effectiveness 4). In contrast, a different participant reported trusting the antigen test result less than PCR because they had heard from family and friends with medical training that antigen tests were not as accurate as PCR (Perceived effectiveness 5).

While participants were generally confident in their own ability to successfully use the at-home antigen test, they were less confident about their co-workers’ abilities, which they believed could undermine the effectiveness of the daily testing intervention. These concerns included not believing that their coworkers would obtain a sufficient swab sample and properly self-administer the test at home (Perceived effectiveness 6), believing that a test administered by a medical professional would be more accurate (Perceived effectiveness 7), and worry that the identity of the test taker and the test result could not be confirmed in a home setting (Perceived effectiveness 8). Finally, some participants raised concerns that a frequent testing regimen would lead to employees increasing risky behavior that could result in more COVID-19 transmission (Opportunity cost 9, 10).

## 4.0 Discussion

As the COVID-19 pandemic continues to evolve with the emergence of new variants (18,19), waning vaccine and natural immunity (20–22), advancements and setbacks in treatment options (23), and changing human behavior in response to these inputs (24,25), new strategies for controlling and adapting to COVID-19 are needed. The Delta and Omicron variants have brought heightened interest in at-home antigen self-testing and the Biden administration has responded with significant resources to rapidly increase the availability of antigen tests in the US beginning late January 2022. The extent to which this intervention will have a significant public health impact remains to be seen but will in part depend on whether, and if so, how, Americans will use these tests. However as of January 2022, to our knowledge no studies have examined the acceptability of self-testing for COVID-19, including serial testing regimens that have been shown most effective at detecting cases.

Our mixed-methods study on the acceptability of a serial testing regimen showed that daily testing using an at-home antigen COVID-19 test was acceptable in one employment context. Employees were willing and able to adhere to a daily testing regimen for up to 21 days, with 75% of participants achieving adherence 90% or greater. In addition to reporting overall high acceptability and adherence, our study identified three key implementation challenges that must be addressed for antigen tests to reach their full public health potential.

First, there is a significant need for educational campaigns to build lay user trust in the accuracy of at-home antigen tests and knowledge around when and how to use them, including how to act on test results (26). Of the seven constructs that comprise acceptability, coherence and perceived effectiveness were most salient and determined whether participants ultimately felt it “was worth it” to test for COVID-19 daily. Importantly with respect to coherence, participants expressed a willingness to tolerate burden (inconvenience, invasiveness), opportunity costs (time), threats to ethicality (loss of privacy) associated with daily testing when they perceived the threat of COVID-19 to be high (high coherence), but indicated that given testing fatigue, this tolerance would not last forever. In the face of declining cases and universal access to vaccination among adults, participants saw a time in the then near future when this testing frequency would no longer be necessary, rendering the intervention less acceptable. As shown in Figure 1, the study was implemented right before the emergence of the Delta variant when case numbers were decreasing, vaccination eligibility was extended to study participants, and the fear of COVID was palpably receding in the public domain and among our study participants. Respondents reported that daily at-home testing made less sense at this moment in the epidemic than it had one month earlier when case numbers were rising and vaccinations were not yet approved. These findings highlight that people’s tolerance for testing intensities change based on their changing perceptions of risk. Users urgently need clear public health guidance around when and how frequently to use at-home antigen tests under different epidemiological conditions, including specific guidance around on- and off-ramps to different testing intensities (27).

We also found that trust in the accuracy of the at-home antigen test was key to whether participants perceived that the intervention would be effective. While most participants reported trusting the results of the test, this trust was primarily grounded in trust in the institutions providing this test and not in scientific evidence supporting test accuracy. Yet given that antigen tests will be distributed and used outside of the employment context, educational campaigns are needed to build trust in the tests, including by increasing lay user knowledge about the accuracy of antigen tests for detecting when users are most infectious and able to transmit to others. Increasing user literacy around antigen testing for COVID-19 may be particularly important given emerging mixed evidence about the performance and accuracy of antigen tests with the Omicron variant, including concerns about tests being less sensitive (28–31) and slower to detect COVID-19 in the nose compared to other sites (32), which may erode trust in antigen tests and impact acceptability and adherence to testing regimens. This work will need to be ongoing and responsive to emerging information with each new variant and delivered across all communities to ensure that scale up antigen testing does not exacerbate existing health disparities (7,33).

Second, we wish to highlight that any frequent testing campaign will generate an enormous amount of sensitive health data and there is currently no existing universal infrastructure for routinely capturing or reporting this information to relevant health authorities, health care providers, or institutions (e.g., employers, schools). The lack of a universal recording and reporting system for at-home antigen test results that are test brand agnostic creates numerous challenges including 1) potentially biased statistics around COVID-19 infection rates due to millions of home antigen test results not being reported, with implications for public health surveillance and resource allocation (34–36); 2) sharing of COVID-19 test results through non-secure channels (e.g., text messages, emails) providing opportunities for privacy breaches (37–39); and 3) challenges for the delivery of anti-viral treatments for COVID-19 within three days after symptom-onset for maximum benefit, due to the lack of guidance on whether a positive home antigen test can be used to qualify for treatment (40,41). In our study, testing data was captured securely and systematically using a mobile app that recorded and interpreted the test result for the user; these data were only shared with the study team, but the app could be further developed to support secure sharing with other entities. Most participants in our study found the app to be acceptable, however using this app was not without challenges, and some participants experienced significant difficulty using the app daily due to lack of internet or smart phone operating system too old to support the app. In a less technology savvy population, comfort with smart phone technology may also be a significant barrier. Further, while nearly all participants were comfortable with tests results being shared with the research team and their employer, they expressed significant concern about results being shared with health or government entities aside from their own provider.

Anticipating potentially more dangerous pandemics in the near future, the Biden administration has developed a five-pillar plan for pandemic preparedness that includes “transforming our medical defense [through] dramatically improving diagnostics” (Pillar 1) and “ensuring situational awareness about infectious disease threats for both early warning and real-time monitoring” (Pillar 2) (42). A connected at-home antigen test that could accurately capture tests results and share them with government and health authorities in real time would address both pillars, however significant work remains to be done to build public trust and acceptability in such a system (43). Within the current pandemic, anecdotally, antigen test results are already being informally shared with institutions like schools and employers to support strategies like “test to stay” (testing to remain after exposure) and “test to return” (testing to end isolation or return after infection). To our knowledge, there has been little to no research done on how these results are currently being shared or stored; guidelines for best practices are urgently needed.

Finally third, given that the COVID-19 and future pandemics may disproportionally and negatively affect low-income and marginalized communities, it is essential that serial testing interventions in the workplace and elsewhere be employed in ways that put equity at the forefront. Mandatory workplace testing policies have the potential to both protect the most vulnerable, front-line employees, but if instituted without proper protections, can also create negative unintended consequences that may disproportionately burden vulnerable populations. The acceptability of a daily testing regimen in this study population was likely influenced by the employment context, including already established norms around routine COVID-19 testing, and job security and benefits (e.g., health insurance, dedicated paid COVID-19 sick leave) available to participants in our study who were all employed full-time. Although we anticipated and probed for worries about job security or loss of income due to testing positive for COVID-19, hypothesizing that this could reduce acceptability and adherence to the intervention, no participants reported such concerns. Possibly because of their job security, participants also almost universally believed that mandatory COVID-19 testing in the workplace was ethical. In other employment contexts with less job and income protection, mandatory serial testing may be much less acceptable and could have harmful unintended consequences.

## 5.0 Conclusion

Our study contributed to the knowledge gap around the implementation of serial at-home rapid antigen testing regimens. This is the first study to our knowledge to evaluate the acceptability of a serial COVID-19 testing regimen with a connected test in an employment context and report on adherence to the intervention. Our study advanced the Theoretical Framework of Acceptability by developing quantitative measures of acceptability and identifying three critical barriers to the successful implementation of rapid antigen testing as a public health intervention.

There were also a number of limitations. The study was implemented at a time of relatively low COVID worry as the number of cases was falling steeply and vaccines offered a promise of durable protection. This epidemiological context along with the COVID-19 vaccine exclusion criterion impacted our ability to enroll a larger number of study participants and limits the generalizability of the findings and our ability to reach saturation. However, low accrual highlighted that context is an understudied component of acceptability causing acceptability to vary across time. Generalizability may also be limited by the fact that the small study was conducted in Southern California in a politically liberal area with high support for COVID precautions and within a stable employment context. Participants were all fully employed at a large company (Disney), and race, education, and SES are not representative of the broader US population. The relatively high adherence to the intervention we report is likely an artifact of the study context and selection bias based on willingness to enroll in the parent daily testing study and participate in in-depth interviews about the experience; we would expect adherence to be lower in the real world. Finally, we only looked at serial testing in an employment setting, although serial testing has now been rolled out in other contexts. Additional studies are needed to build the evidence-base about the acceptability of and adherence to serial-testing regimens to evaluate their public health contribution to interrupting transmission.

## Data Availability

The data cannot be shared publicly because of ethical restrictions and data protection issues as our dataset includes potentially identifying information.

